# Change for life? Adolescent cognitive development predicts mortality risk independent of childhood ability

**DOI:** 10.64898/2026.05.23.26353598

**Authors:** Kristine B. Walhovd, Anne Ingeborg Berg, Sandra Buratti, Jonas Burén, Pär Bjälkebring, Martin Fischer, Isabelle Hansson, Linda Hassing, Anna-Carin Jonsson, Lina Jonsson, Magnus Lindwall, Therese Nilsson, Ole Røgeberg, Andreas Segerberg, Valgeir Thorvaldsson, Mikael Landén, Alli Klapp, Martin Lövdén

## Abstract

Lower cognitive ability measured in childhood or late adolescence has been consistently associated with higher mortality risk across adulthood. However, this evidence largely relies on single assessments, leaving it unclear to what extent mortality risk reflects cognitive differences established early in life versus developmental divergence during adolescence—a period of substantial neurocognitive plasticity. Using two nationally representative Swedish cohorts comprising 9,412 males born in 1948 and 1953, we linked cognitive ability assessed in primary school at age 13 years and military conscription at age 18 years to all-cause and cause-specific mortality recorded in nationwide registers through 2025. We decomposed late-adolescent cognitive ability into childhood cognitive level and adolescent cognitive change and evaluated their independent associations with mortality. Childhood cognitive level (HR = 0.81; 95% CI, 0.78–0.85) and adolescent cognitive change (HR = 0.84; 95% CI, 0.79–0.89) independently predicted lower mortality risk, also after adjustment for parental education. Childhood cognitive level and adolescent cognitive change showed partially distinct cause-specific patterns. Childhood cognitive level was most strongly associated with mortality from intrinsic causes, whereas adolescent cognitive change showed relatively stronger associations with external causes, particularly accidental deaths. Although adolescent cognitive change was associated with psychosocial factors including education and psychiatric diagnosis at conscription, its association with mortality persisted after adjustment for these factors. These findings suggest that cognitive development during adolescence carries independent prognostic information regarding long-term survival beyond cognitive level established by late childhood, highlighting adolescence as a consequential period for lifelong health.

## Introduction

Cognitive ability measured in childhood or early adulthood is robustly associated with subsequent mortality risk, with lower scores predicting higher all-cause and cause-specific mortality across decades of follow-up (1–6). These associations have been replicated across cohorts, countries, and historical periods, establishing early cognitive ability as one of the strongest psychosocial predictors of longevity.

Despite this consistency, most studies have relied on cognitive assessment at a single time point. Whether measured in childhood or at military conscription, cognitive ability in youth is typically treated as relatively stable, implicitly assuming that developmental change prior to adulthood contributes little additional information regarding long-term health outcomes. This assumption contrasts with evidence from developmental psychology and cognitive neuroscience showing that adolescence is characterized by substantial neurobiological, cognitive, and psychosocial development (7–10). Cognitive abilities continue to improve on average through adolescence, while rank-order stability remains incomplete, allowing meaningful individual differences in developmental trajectories. Adolescents with similar childhood cognitive levels may therefore diverge by late adolescence.

In contrast to the lack of knowledge on the predictive value of development, cognitive change has been extensively studied in midlife and older adulthood, where accelerated cognitive decline independently of baseline level is strongly associated with increased mortality risk (11–15). Although these findings concern aging-related decline, they suggest more broadly that developmental trajectories, rather than cognitive level alone, may carry prognostic information. Indirect support also comes from studies of social and educational mobility showing that changes across the life course predict mortality beyond childhood socioeconomic conditions (16–18). More directly, a small number of studies have examined developmental cognitive trajectories in relation to later outcomes, including long-term mortality and suicide risk (19–23). However, these studies have focused on specific outcomes or developmental indicators and have not directly examined whether cognitive developmental divergence during adolescence independently predicts mortality across major causes of death.

Recent work in longevity research has renewed interest in distinctions between intrinsic and extrinsic mortality processes (24, 25). External causes of death such as accidents and violence may reflect developmental and behavioral pathways distinct from those contributing to chronic disease and aging processes (25–29). Childhood and adolescent cognitive difficulties have previously been linked to higher risk of suicide and accidental death (21–23) raising the possibility that developmental differences during adolescence may be especially relevant for mortality arising from external causes.

Here, we examine whether cognitive development across adolescence predicts mortality risk independently of childhood cognitive level. Using two nationally representative Swedish cohorts followed across adulthood, we decomposed late-adolescent cognitive ability into childhood cognitive level and adolescent cognitive change and examined their independent associations with all-cause and cause-specific mortality. We further explored whether childhood cognitive level and adolescent developmental divergence showed different patterns across intrinsic and external causes of death.

## Results

### Study sample and follow-up

The analytic sample comprised 9,412 men from two nationally representative Swedish birth cohorts (UGU 1948 and 1953 (30, 31)), analyzed jointly using cohort-stratified baseline hazards. Participants were included if they had complete cognitive data at age 13 years (primary school testing (30)) and age 18 years (military conscription (32)), including all subtests contributing to the general cognitive ability measures at both assessments, as well as parental education recorded in childhood. Participants were followed for mortality through linkage with the nationwide Cause of Death Register until November 3, 2025. During follow-up, 2,247 deaths occurred between 1967 and 2025. The median attained age at death was 65.9 years (mean = 61.6, SD = 13.5, range = 18.5 - 77.6). Characteristics of the analytic sample is given in Table 1, see SI for further information on sampling frame.

**Table 1.** Sample characteristics. Values are presented as n for categorical variables and mean (SD) for continuous variables. Follow-up time is calculated from age 18 years to death or administrative censoring (November 3, 2025). Follow-up differs by survival status because deceased individuals contribute person-time until death. Cognitive ability was extracted as latent g, where childhood cognitive ability is given in z-scores (mean = 0, SD+/-1)- Adolescent cognitive change represents late-adolescent cognitive ability residualized with respect to childhood cognitive level.

| Characteristic | Alive | Dead |
| --- | --- | --- |
| N | 7165 | 2247 |
| 1948 cohort, n | 3585 | 1416 |
| 1953 cohort, n | 3580 | 831 |
| Childhood cognitive level, mean (SD) | 0.06 (0.99) | -0.19 (1.00) |
| Adolescent cognitive ability, mean (SD) | 0.07 (0.98) | -0.22 (1.03) |
| Adolescent cognitive change (residual), mean (SD) | 0.02 (0.63) | -0.07 (0.66) |
| Parental education, mean (SD) | 0.02 (1.02) | -0.10 (0.85) |
| Age at death, mean (SD), y | — | 61.64 (13.50) |
| Follow-up time, mean (SD), y | 56.86 (2.52) | 43.64 (13.50) |

### Cognitive measures and developmental modeling

Cognitive ability at ages 13 and 18 was derived from multiple subtests and summarized as a latent general cognitive ability factor (g) at each wave. Scores were standardized (z-scores) within wave, such that a one-unit difference corresponds to one standard deviation within the cohort at that age. Childhood and adolescent g were strongly correlated (r = .77), indicating substantial stability in cognitive performance across adolescence while allowing for meaningful individual differences in developmental divergence. Adolescent cognitive change was operationalized as late-adolescent performance relative to earlier standing, quantified as the residual of age-18 g after regressing on age-13 g. Positive values indicate higher-than-expected performance at age 18 given childhood ability; negative values indicate lower-than-expected performance; zero represents the sample-average developmental trajectory (Figure 1). Because age-18 g was standardized (z-scored) prior to residualizing, the residual retains the metric of age-18 g: one unit of the residual corresponds to one SD of late-adolescent g, net of childhood level. Effect sizes for childhood level and adolescent change are therefore directly comparable, as both reflect mortality risk per one SD of g at their respective ages. The SD of the residual (∼0.64) reflects the realistic constraint that developmental divergence across adolescence is attenuated relative to the full distribution of age-18 g; re-standardizing the residual would remove this property and render the two predictors non-comparable in g units.

**Figure 1.**
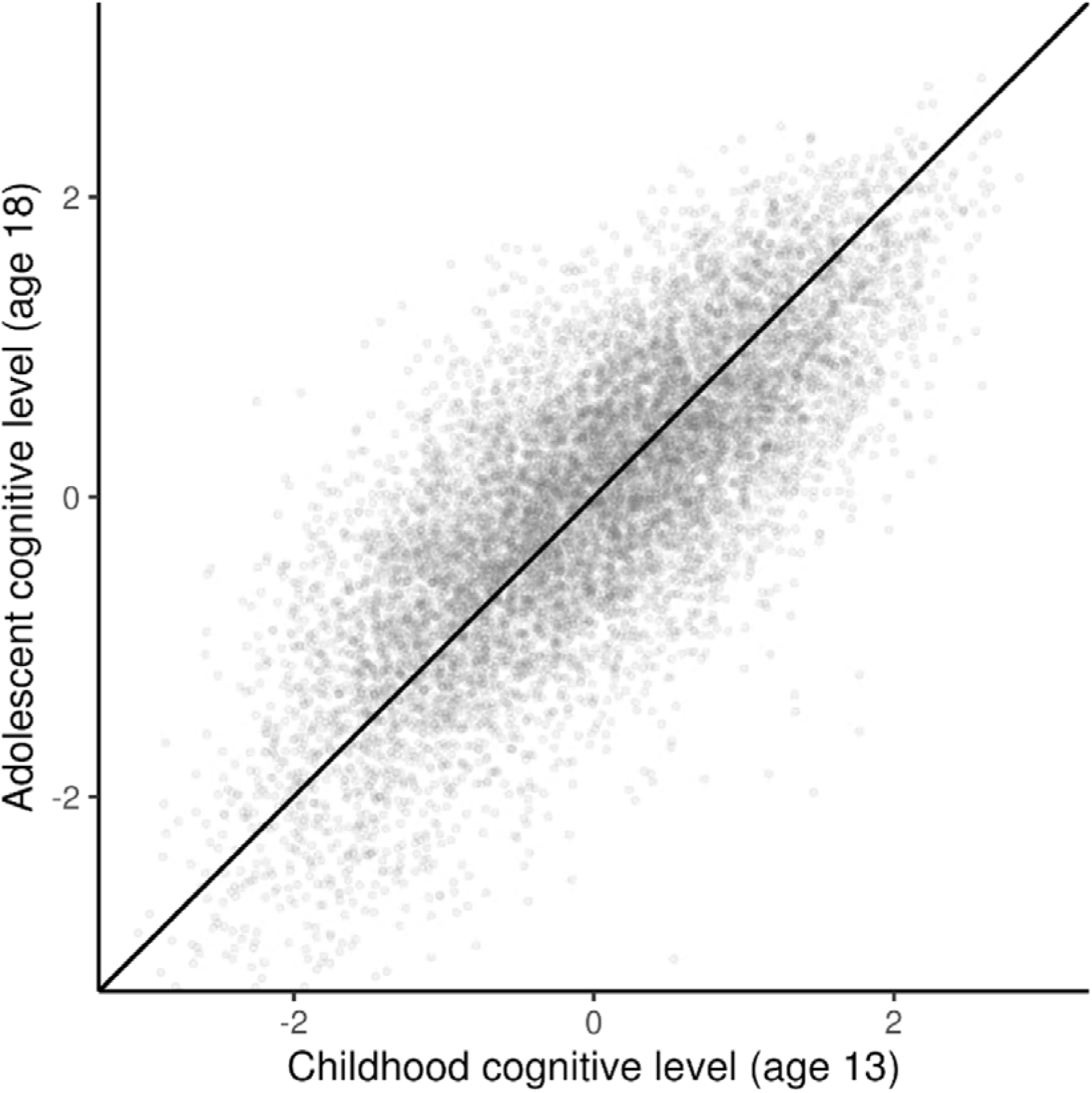
Developmental divergence in cognitive ability from childhood to adolescence. Childhood cognitive level at age 13 plotted against adolescent cognitive level at age 18 (z-standardized). The identity line (slope = 1) represents no developmental change. Dispersion around this line reflects individual differences in cognitive development across adolescence.

By construction, the residualized change measure is uncorrelated with childhood cognitive ability, allowing their independent contributions to mortality risk to be estimated. Results were similar using simple difference scores (Δg; SI), suggesting that findings were not highly dependent on the specific operationalization of adolescent developmental divergence. We used residualized adolescent cognition in main analyses rather than raw difference scores because residualization isolates cognitive information at age 18 beyond childhood level while avoiding some of the reliability limitations of raw change scores when repeated measures are strongly correlated. However, under classical measurement error, residualized scores may still partly reflect stable trait variance not captured by the childhood measure rather than genuine developmental change. We therefore conducted supplementary measurement-error calibrated analyses using externally anchored estimates of latent cognitive stability (33) to evaluate the extent to which the residualized measure reflected developmental divergence versus recovery of stable trait variance (see SI, Decomposition and measurement-error calibration of the residualized adolescent cognitive measure).

### Cognitive ability in childhood and late adolescence and mortality risk

Using Cox proportional hazards models with attained age as the time scale and cohort-stratified baseline hazards, we first examined childhood and adolescent cognitive ability separately. One standard deviation higher childhood cognitive ability at age 13 was associated with 20% lower mortality risk across adulthood (hazard ratio (HR) per SD = 0.80; 95% CI, 0.77–0.83). Cognitive ability at age 18 showed a similar association when modeled alone (HR per SD = 0.78; 95% CI, 0.75–0.81).

### Childhood cognitive level and adolescent cognitive change independently predict adult mortality

We next examined whether adolescent cognitive change – defined as performance at age 18 relative to childhood level – was associated with mortality beyond childhood cognitive ability. When modeled jointly, both components were independently associated with mortality risk. One-standard deviation higher childhood cognitive level was still associated with 20% lower mortality risk (HR = 0.80; 95% CI, 0.77–0.83), while a one standard deviation more positive adolescent cognitive change was associated with an additional 17% lower risk (HR = 0.83; 95% CI, 0.78–0.88). Adjustment for parental education did not alter this pattern: childhood cognitive level (HR = 0.81; 95% CI, 0.78–0.85), and adolescent cognitive change (HR = 0.84; 95% CI, 0.79–0.89) remained independently associated with lower mortality risk. Higher parental education was more modestly associated with lower mortality risk (HR per SD = 0.95; 95% CI, 0.90–0.996) (Figure 2). Estimates using non-residualized measures are reported in the SI.

**Figure 2.**
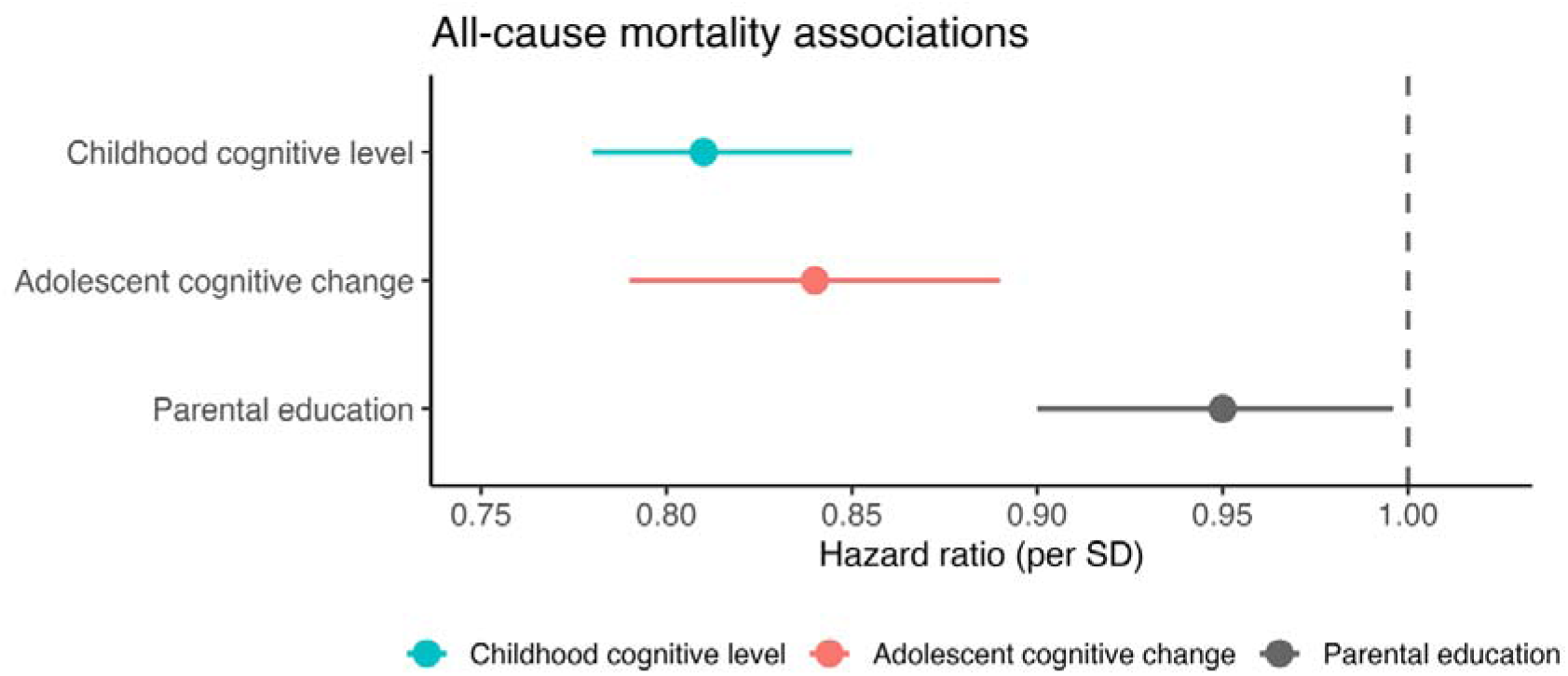
Childhood cognitive level and adolescent cognitive change in relation to all-cause mortality. Panel Hazard ratios (HRs) and 95% confidence intervals for all-cause mortality per 1-SD higher childhood cognitive level at age 13 years (child g), adolescent cognitive change from childhood to late adolescence (army g residual), and parental education. Estimates are from Cox proportional hazards models with attained age as the time scale, stratified by birth cohort and mutually adjusted for all variables shown. HRs < 1 indicate lower mortality risk.

These results show that mortality risk is independently associated with both stable differences in cognitive level established by early adolescence and individual differences in cognitive development between ages 13 and 18 years.

### Cause-specific mortality predicted by childhood level and adolescent change

To examine whether associations differed across causes of death, we estimated cause-specific Cox models including childhood cognitive level and adolescent cognitive change simultaneously (Figure 3; Table 2). Distinct patterns emerged for intrinsic and external causes. For major disease categories, childhood cognitive level showed the most consistent associations with mortality. Higher childhood cognitive level was associated with lower mortality from cancer, cardiovascular disease, and respiratory disease. In contrast, adolescent cognitive change tended to show weaker or less consistent associations with these outcomes. Cancer mortality showed no clear association with adolescent cognitive change, whereas cardiovascular mortality was associated with both childhood level and adolescent change to a similar extent.

**Figure 3.**
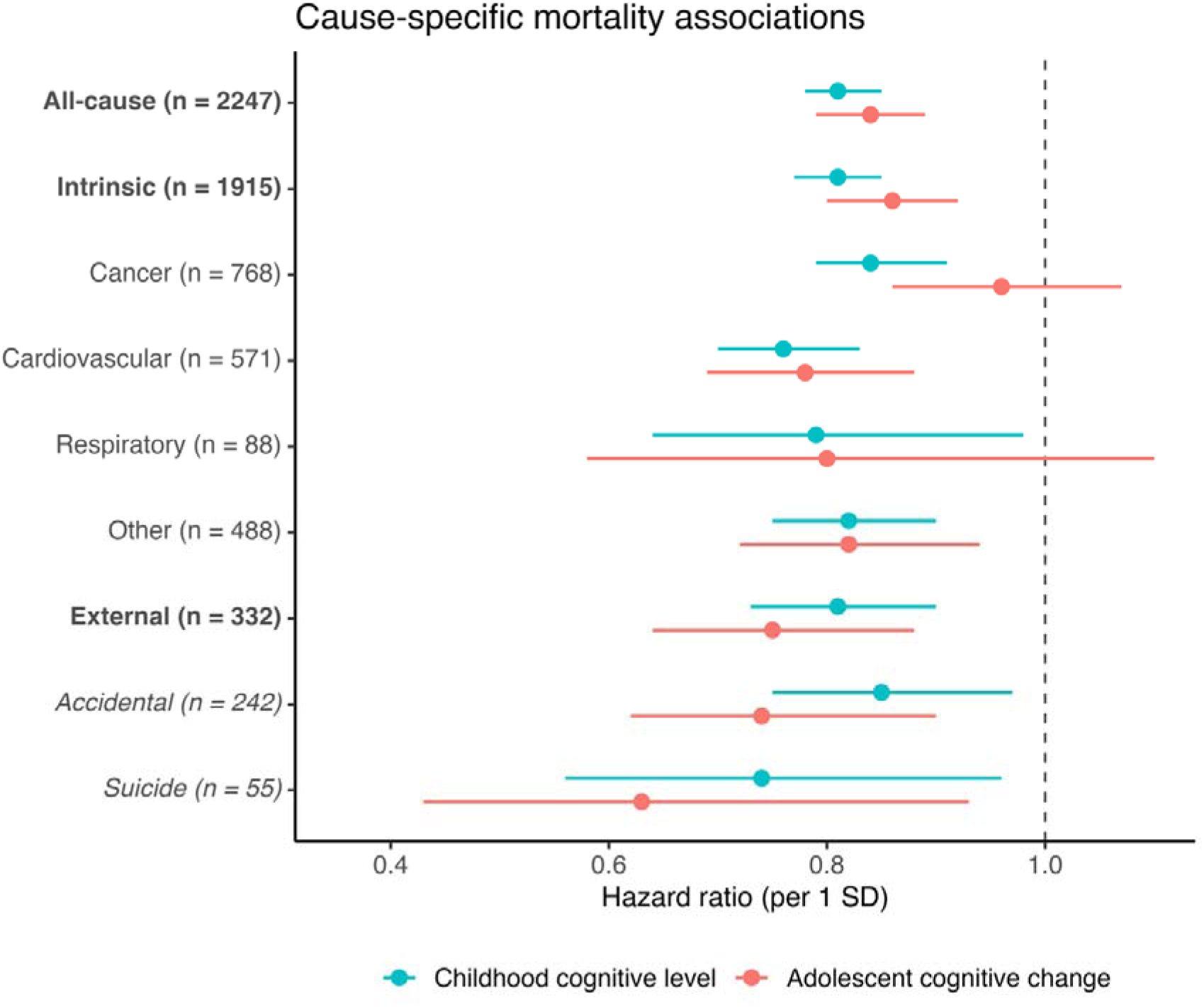
Cause-specific mortality: childhood cognitive level and adolescent cognitive change. Hazard ratios (HRs) with 95% confidence intervals (CIs) from Cox proportional hazards models estimating associations of childhood cognitive level (age 13) and adolescent cognitive change (residualized cognitive ability at age 18, adjusted for age-13 level) with cause-specific mortality. Models include both components simultaneously and are adjusted for parental education. Estimates are shown per 1–standard deviation increase in each predictor. Intrinsic deaths include cancer, cardiovascular disease, respiratory disease, and “other”. External causes accidental and suicide are shown, while homicides (n = 24) and undermined intent (n =11) are not shown here. Details are reported in Table 2.

**Table 2.** Cause-specific mortality associations with childhood cognitive level and adolescent cognitive change. Hazard ratios (HR) and 95% confidence intervals from cause-specific Cox proportional hazards models with attained age as the timescale, adjusted for birth cohort and parental education. Childhood cognitive level and adolescent cognitive change were entered simultaneously in each model. Adolescent cognitive change is operationalized as the residual of age-18 g after regressing on age-13 g; one unit corresponds to one SD of late-adolescent g net of childhood level (see Methods). Deaths from undetermined causes (n=11) and homicide (n=24) are shown for completeness but should be interpreted with caution given sparse events. External causes include accidental death, suicide, homicide, and undetermined causes, and subsets of external causes are shown separately given their theoretical relevance to intrinsic versus extrinsic mortality.

| **Cause** | **Deaths** | **Predictor** | **HR (95% CI)** | **p** |
| --- | --- | --- | --- | --- |
| **Intrinsic** | 1915 | Child level | 0.81 (0.77–0.85) | <0.001 |
|  |  | Adolescent change | 0.86 (0.80–0.92) | <0.001 |
| Cancer | 768 | Child level | 0.84 (0.79–0.91) | <0.001 |
|  |  | Adolescent change | 0.96 (0.86–1.07) | 0.446 |
| Cardiovascular | 571 | Child level | 0.76 (0.70–0.83) | <0.001 |
|  |  | Adolescent change | 0.78 (0.69–0.88) | <0.001 |
| Respiratory | 88 | Child level | 0.79 (0.64–0.98) | 0.035 |
|  |  | Adolescent change | 0.80 (0.58–1.10) | 0.167 |
| Other | 488 | Child level | 0.82 (0.75–0.90) | <0.001 |
|  |  | Adolescent change | 0.82 (0.72–0.94) | 0.004 |
| **External** | 332 | Child level | 0.81 (0.73–0.90) | <0.001 |
|  |  | Adolescent change | 0.75 (0.64–0.88) | <0.001 |
| *Accidental* | 242 | Child level | 0.85 (0.75–0.97) | 0.015 |
|  |  | Adolescent change | 0.74 (0.62–0.90) | 0.002 |
| *Suicide* | 55 | Child level | 0.74 (0.56–0.96) | 0.024 |
|  |  | Adolescent change | 0.63 (0.43–0.93) | 0.019 |
| *Homicide* | 24 | Child level | 0.53 (0.35–0.80) | 0.003 |
|  |  | Adolescent change | 0.98 (0.54–1.76) | 0.944 |
| *Undetermined* | 11 | Child level | 1.06 (0.57–1.97) | 0.848 |
|  |  | Adolescent change | 1.23 (0.48–3.17) | 0.669 |

For external causes of death, adolescent cognitive change contributed more strongly. Both childhood cognitive level and adolescent cognitive change were associated with lower mortality from external causes overall, but the association with adolescent change was larger. This pattern was most evident for accidental deaths, where more positive adolescent cognitive change was more strongly associated with lower mortality risk than childhood cognitive level. For suicide, both components were associated with mortality risk, and even though adolescent cognitive change here appeared more strongly related, cases are few, and estimates were less precise, so caution is warranted in the interpretation of these estimates (see below). Estimates for homicide and deaths of undetermined intent were imprecise because of low numbers of events and are shown for completeness but should be interpreted cautiously. Overall, childhood cognitive level showed the strongest and most consistent associations with mortality from intrinsic causes, especially cancer, whereas adolescent cognitive change showed relatively stronger associations with external causes, particularly accidental mortality.

To evaluate whether these descriptive differences reflected coefficient heterogeneity, we conducted an exploratory formal comparison contrasting cancer and accidental mortality. Associations of adolescent cognitive change differed across these causes (p = .020), indicating a relatively stronger association with accidental than cancer mortality. In contrast, associations of childhood cognitive level did not differ across causes (p = .78). The joint test of both cognitive predictors showed a trend-level difference across mortality categories (p = .062).

### Measurement-error calibrated estimates

Under the externally anchored measurement model, approximately 92% of the non-noise cognitive signal in the residualized adolescent score was estimated to reflect genuine adolescent change, with approximately 8% reflecting stable-trait variance not captured by the childhood test (Figure S2). The genuine-change share stays in the 86%-92% range across the CI of the external anchor, yielding adjusted HRs on the cognitive change for all-cause mortality in the 0.79-0.84 range. About half of the total residualized-score variance was estimated to reflect measurement noise, implying attenuation of the observed Cox coefficient. Regression-calibrated Cox estimates showed the same overall pattern as the primary observed-score analyses but sharpened the cause-specific contrast: the weak adolescent-change association for cancer mortality was further attenuated, whereas associations with external causes, especially accidental mortality, strengthened (Figure S3). These results support the interpretation of the residualized adolescent score as primarily developmental divergence rather than recovery of stable trait variance, while retaining the observed-score Cox estimates as the primary analyses.

### Correlates of adolescent cognitive change and adjustment

To contextualize what adolescent developmental divergence may capture, we explored available educational, psychosocial, and health-related correlates of adolescent cognitive change within each cohort (variables varied by cohort; see Methods and SI). In the 1948 cohort, longer distance to school beyond primary education and having a father who was a farmer were associated with less positive cognitive change. In the 1953 cohort, educational attainment at conscription was strongly positively associated with cognitive change, whereas psychiatric morbidity, particularly substance-related disorders, and to some extent reduced eyesight, were associated with less positive change. Across both cohorts, higher parental education was consistently associated with more positive cognitive change (Table S1). These findings suggest that individual differences in adolescent cognitive development partly reflect early-life environmental conditions and educational pathways. The present study was not powered to formally test by which factors the relationship of adolescent cognitive change and mortality risk are mediated. Exploratory adjustment for these factors did not substantially attenuate the relationship between adolescent cognitive change and all-cause mortality in either cohort. In the 1948 cohort, none of these variables were significantly associated with all-cause mortality in this model. In the 1953 cohort, psychiatric and substance abuse diagnoses at conscription were associated with increased all-cause mortality risk. For accidental mortality, introduction of these covariates did not substantially attenuate the associations of child cognitive level and adolescent cognitive change in the 1948 cohort, while they were somewhat weakened and reduced non-significant for child level and to a trend for cognitive change when including these covariates for the 1953 cohort, with a trend for substance abuse diagnosis at conscription to increase risk of accidental death (SI, Tables S2–S5).

## Discussion

This population-based study shows that cognitive development across adolescence predicts adult mortality independently of cognitive level established in late childhood. By decomposing late-adolescent cognitive ability into childhood cognitive level and adolescent change, we found that these components relate to mortality in partly distinct ways across adulthood and causes of death. Despite substantial stability in individual differences in cognitive ability from ages 13 to 18, individual differences in change were associated with later mortality risk, with the clearest associations observed for external causes, especially accidental deaths – which dominate mortality in young adulthood. These findings advance a literature previously mostly reliant on single early-life cognitive assessments (1, 2, 5, 34, 35). It is also consistent with previous studies linking developmental problems more strongly to death from extrinsic than intrinsic causes (22). By disentangling childhood cognitive level and adolescent cognitive development, we demonstrate that the latter carries distinct prognostic information.

Cause-specific analyses suggested partly distinct patterns for childhood cognitive level and adolescent cognitive change. Childhood cognitive level tended to be more strongly related to mortality from major disease categories, particularly cancer, while both childhood level and adolescent cognitive change were associated with cardiovascular mortality. Associations of cognitive function at young age are consistent with prior work for cardiovascular disorders (5, 34, 36). In contrast to low associations with cancer mortality, adolescent cognitive change tended to be more strongly associated with external causes of death, especially accidental deaths, which are proportionately more common at younger adult ages. These patterns thus further suggest that cognitive development across adolescence may capture processes distinct from those reflected by stable cognitive differences established earlier in life (37), and point to the possibility that these processes may be particularly relevant for mortality risks relatively more common in early adulthood. The present study is not sufficiently powered to definitively identify differential associations for specific smaller classes of death causes. However, exploratory formal heterogeneity analyses further supported this interpretation in comparisons of the largest contrasted intrinsic and external mortality categories. In analyses contrasting cancer and accidental mortality, associations of adolescent cognitive change differed across causes, with stronger associations observed for accidental than cancer mortality, whereas childhood cognitive level showed no corresponding difference. Although exploratory and based on selected cause contrasts, these findings paralleled the descriptive cause-specific patterns and are consistent with the possibility that developmental divergence across adolescence may be particularly relevant for mortality processes linked to behavioral and environmental risk exposure.

Several explanations have been proposed for associations between early-life cognitive ability and mortality, including early health and social disadvantage, health literacy and self-regulation, and broader system-integrity accounts in which cognitive performance indexes general physiological robustness (4, 36). These frameworks may help explain why lower cognitive ability in youth predicts both disease and external-cause mortality in prior studies (2, 3, 38–41). However, they translate less directly to adolescent cognitive change. One interpretation is that developmental divergence during adolescence captures at least one of two processes: developmental processes relevant to later behavioral regulation, educational sorting, or exposure to hazardous environments (2–4, 42–44); or, alternatively, that it partly reflects subclinical health or neurodevelopmental liabilities emerging during adolescence (45).

Crucially, the association between adolescent cognitive change and external mortality, especially accidental death, cannot be interpreted as evidence that these outcomes are fixed consequences of individual traits. On the contrary, external deaths are by definition highly contingent on context, exposure, and opportunity for harm. A more cautious interpretation is that adolescent cognitive development may index vulnerability or resilience in processes relevant to the navigation of risk, including educational placement, self-regulation, problem-solving, and exposure to hazardous social or physical environments (43). This interpretation is consistent with developmental research showing that adolescence is marked by continuing maturation of cognitive control, changing sensitivity to reward and peers, and a high burden of injury in youth and young adulthood (44).

To interrogate these possibilities, and to test whether these effects are byproduct of structural disadvantages, our follow-up analyses examined associations with both socio-educational and health-related proxies. As expected, adolescent cognitive trajectories were related to both domains. Positive cognitive change was associated with socio-environmental opportunity, indexed by factors such as parental and own education, which is tied to genetic factors that may directly or indirectly influencing cognitive function (46, 47). Relatively negative cognitive change associated with geographic distance to non-compulsory schooling, having a farmer background, emergent health vulnerabilities, particularly psychiatric symptoms, substance-related disorder and poorer eyesight. We are not, with the observed effects sizes and the current sample size, powered to determine whether such factors mediate mortality risk. However, adjustment for these factors did not substantially attenuate associations of adolescent cognitive change and accidental mortality in the 1948 cohort, while in the 1953 cohort, the inclusion of these variables rendered the association weakened to a trend, with a trend for substance abuse diagnosis at conscription to associate with accidental death. In all-cause models, associations of both child level and adolescent cognitive change remained significant also when including other psychosocial and diagnostic predictors, even if psychiatric and substance abuse diagnose were associated with increased mortality risk. This may indicate that adolescent cognitive development is not merely a downstream proxy for overt socioeconomic disadvantage or psychiatric illness (45, 48, 49). Rather, it may capture distinct developmental signals, such as the above-mentioned shifts in cognitive control and risk navigation, with enduring consequences for later-life survival.

Ultimately, these developmental and behavioral dynamics may map onto longstanding conceptual and empirical parsing of mortality into biologically meaningful categories (28, 29), that has recently regained attention in longevity research (24, 25), namely the distinction between intrinsic and extrinsic mortality processes. Recent work has shown that deaths from external causes, such as accidents, violence, and intoxication, essentially mask the observable heritability of longevity by conflating intrinsic biological disease and aging with extrinsic mortality processes (25). When extrinsic mortality is accounted for, the heritability of intrinsic human lifespan is substantially higher, exceeding 50%. Within this framework, these results may suggest that adolescent cognitive development may be the most linked to mortality processes that are extrinsic in nature, whereas childhood cognitive level may most strongly differentiate risks related to disease processes that unfold across adulthood. In this sense, cognitive development across adolescence may index vulnerability – or resilience – to environmental, behavioral, and social risks that dominate early adult mortality (43, 44), rather than intrinsic disease or aging processes per se (25, 28, 29) causing deaths concentrated at later life stages (see Figure S1). This is also supported by adolescent cognitive change being significantly more predictive of accidental death than of cancer.

The observed associations of adolescent cognitive change and external, especially accidental causes of death, likely have several, non-exclusive explanations. First, external causes of death are concentrated in earlier adulthood. Pronounced rises in mortality rates are seen from early adolescence to young adulthood, with external causes dominating (26), so developmental differences relevant to those outcomes may matter most when those exposures are most common. Second, selective survival may reduce observable differences at older ages, because individuals at highest risk have already died (4). Third, as chronic disease burden accumulates across adulthood, mortality may increasingly reflect later-life exposures and biological aging processes not well indexed by cognitive differences measured in youth (50).

The association between young cognitive ability and cancer mortality has been inconsistent in prior studies (35, 42, 51). Large-scale analyses of Swedish conscripts have reported no association between early adult cognitive ability and cancer mortality by midlife, while other cohorts have shown weak or cause-specific associations. One element that differs between some of these analyses (42) and the present, beyond historical changes and developmental timing of measurement, is that that the current cohorts were born within a relatively narrow interval (∼5 years) and followed into their 70s. Because cancer deaths are heavily concentrated at older ages, analyses that focus on varying follow-up periods, including shorter age ranges, may not capture to the same extent cognitive differences may be more relevant for cancer deaths across the lifespan. Prior work also suggests that associations may be concentrated in smoking-related and digestive cancers (2, 35). The present study used a broad cancer category. Accordingly, the observed association between childhood cognitive level and cancer mortality should not be interpreted as evidence for a general relation between childhood cognition and cancer biology. It may instead reflect a mixture of cancer-site composition, smoking and other health behaviors, socioeconomic pathways, and survival or detection processes.

### Reliability-bounded interpretations

An important consideration is that cognitive ability was assessed using different test batteries at ages 13 and 18, precluding direct interpretation of absolute change on a common scale. Accordingly, adolescent cognitive change in the present study reflects relative change in standing within the cohort – i.e., whether individuals perform better or worse in late adolescence than expected based on their childhood level – rather than absolute developmental gain or decline.

A central interpretive issue is still whether the residualized adolescent cognitive measure primarily reflects genuine developmental divergence or artefacts of measurement error. Under classical measurement error, residualized scores can partly capture stable trait variance not measured reliably at baseline. However, supplementary externally anchored (33) measurement-error analyses suggested that most of the cognitive signal in the residualized measure reflected genuine adolescent change rather than recovery of stable trait variance. Moreover, regression-calibrated estimates strengthened rather than weakened the observed associations for external and accidental mortality, while the already weak association with cancer mortality attenuated further. Although these calibrated estimates depend on assumptions regarding latent stability and measurement structure, they support interpreting adolescent cognitive change as a meaningful developmental construct rather than a statistical artefact. This rank-based operationalization thus appears to provide a robust and interpretable estimate of developmental differences given the absence of measurement invariance across ages.

### Limitations

Several limitations should be noted. The sample comprised men only, limiting generalization to women. This limitation is especially noteworthy regarding the findings on accidental death, as males are substantially more likely to die in accidents (52), and we must emphasize that these findings do not necessarily apply to females. The UGU cohort is drawn from a Swedish population-representative birth cohort, and data on ethnicity specifically were not collected. Generalizability to more diverse populations is uncertain. Next, although the study was designed to be as population representative as possible, neither the childhood nor the older adolescence testing covers the full population of males. Especially, conscription testing is voided for certain diseases with relevance to mortality (32), and although modest positive selection effects of childhood cognitive levels was observed for conscription testing (see SI), this constitutes a limitation in coverage. The age-13 and age-18 cognitive batteries were overlapping but not identical, and the residualized change measure should be interpreted as developmental divergence in general cognitive performance relative to peers. As with all residualized change measures, it captures variance unexplained by childhood level and is therefore informative but not mechanistically specific. Cause-of-death categories were harmonized across ICD revisions, which is necessary in long follow-up but may introduce some classification uncertainty. Finally, although adjustment for parental education and the available adolescent educational and health variables did not explain the relations of adolescent cognitive change and mortality, we were underpowered for this type of analysis, and residual confounding by family, genetic, behavioral, or environmental factors remains likely (3, 4). Importantly, while the present study points to possible relative different patterns of predictivity for adolescent cognitive change and childhood level across causes of death, we were not powered to do fine-grained analyses of this and the findings should ideally be replicated in future studies. The assignment of cognitive childhood level to age 13 is based on available data, and it may be that also childhood trajectories prior to this can carry predictive information beyond earlier developmental level.

### Conclusions

The mortality relevance of cognitive function in youth is not exhausted by childhood level alone. Developmental change across adolescence carried independent prognostic information. This appeared most evident for external, especially accidental, mortality occurring earlier in adulthood. These findings argue for a life-course perspective in which cognitive development during adolescence is not merely a transition toward adult ability, but part of the developmental architecture through which later health and survival are differentiated. Understanding the factors that shape cognitive trajectories during this period may therefore provide important insights into how developmental processes in adolescence contribute to lifelong differences in health and survival.

## Methods

Approval for the study was given by the Swedish Ethical Review Authority (case number 2024-06028).

## Sample

The UGU study is a Swedish longitudinal database of nationally representative samples of students from multiple cohorts, starting in 1961 (31, 53). Here we include all children of the 1948 and 1953 birth cohorts who could have CDR registry outcomes (see SI, Figure S4) and who lived until at least 18 years of age and with complete data for cognitive tests in 6^th^ grade (1961/1966) and at army conscription age 18 (1948 cohort: n = 5001; 1953 cohort: 4411 males) and parental education at first testing in the analysis. Of these, 2,247 (23,9%) had died as recorded in the Cause of Death Register (CDR), per November 3rd 2025. See SI for details on parental education and other psychosocial measures, as well as analyses on sample selectivity and possible loss to follow-up.

## Cognitive assessments, latent g and residualized change

At age 13 years, participants completed a battery of standardized cognitive tests administered in school. A general cognitive ability factor (child g) was derived and standardized within cohort. At military conscription (≈18 years), participants completed a comprehensive cognitive test battery. A general cognitive ability factor (army g) was derived and standardized within cohort. Because different cognitive test batteries were administered at ages 13 and 18, scores are not directly comparable on a common scale. To separate baseline cognitive ability from developmental differences across adolescence, we decomposed adolescent cognitive ability into: childhood cognitive level (child g), and adolescent cognitive change, operationalized as the residual from a linear regression of army g on child g (army g residual). This measure captures individual differences in cognitive development relative to expected rank-order stability, that is, whether an individual performs better or worse at age 18 than expected given their childhood level, rather than absolute change in cognitive ability. Positive residual values indicate higher-than-expected adolescent performance given childhood ability, whereas negative values indicate lower-than-expected performance. We performed additional measurement-error calibrated sensitivity analyses to take into account differences in error in the cognitive measures used (see Statistical Analyses and SI).

## Cognitive tests

While the specific cognitive subtests administered at ages 13 and 18 were not identical, both batteries contained overlapping domains of cognitive ability, namely logical reasoning, verbal and spatial ability. Notably, test scores of both batteries have been reported to have acceptable, and similar reliabilities, around .90 (range: .87-.92) (30, 54), supporting longitudinal analyses of cognitive level and change. Score distributions for the two cohorts at age 13 and 18 years are shown in Figure 4.

**Figure 4.**
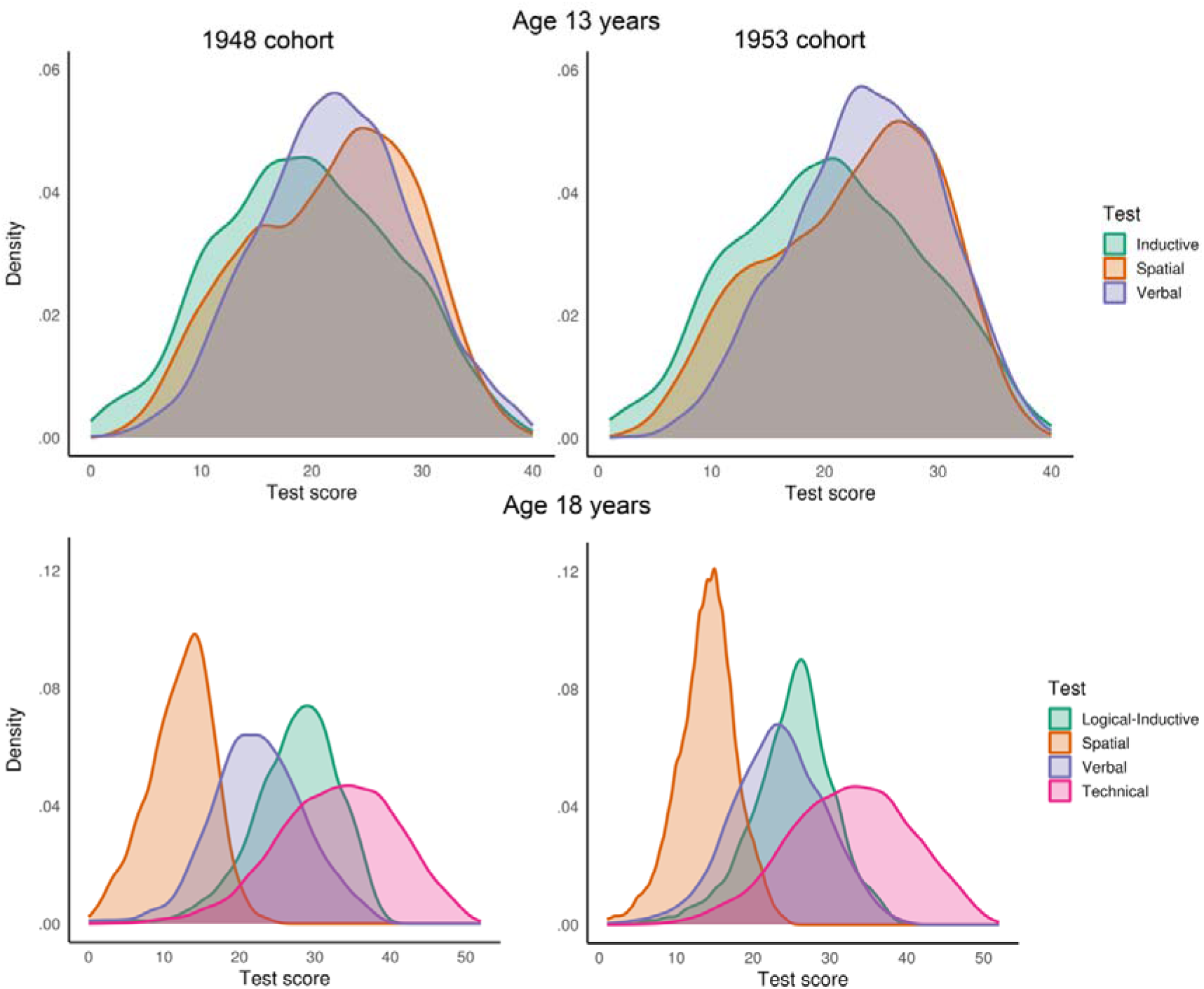
Distributions of general cognitive ability in childhood and late adolescence. Kernel density distributions of standardized general cognitive ability scores at age 13 years (child g) and at age 18 years (army g), shown for the pooled analytic sample. Despite substantial overlap and high rank-order stability between childhood and late-adolescent cognitive performance, distributions indicate meaningful individual differences in cognitive development across adolescence. These individual differences form the basis for the residualized adolescent cognitive change measure used in subsequent analyses. Distributions were similar across cohorts (1948 and 1953), supporting pooled analyses with cohort-stratified baseline hazards.

### Age 13 test battery

In primary school, verbal, spatial abilities and inductive reasoning were assessed with three subtests: 1) antonyms (choosing the opposite of a word from four options), 2) metal folding (determine which of four figures you get if you fold a pictured “sheet metal piece”), and 3) number series (continuing a number-series, where six numbers are given, with two more numbers (30).

### Age 18 test battery

At army conscription, verbal, spatial, reasoning and technical abilities were assessed with four subtests (32, 54, 55): *Instructions* (Logical) was used to measure logical-Inductive ability, and the task was to solve complex verbal instructions and logical puzzles, often requiring following a set of rules to find a specific outcome. *Concept Discrimination* was used to measure verbal ability/comprehension, and the tasks consisted of identifying which word in a set does not belong or selecting a synonym/antonym. *Paper Form Board* was used to test visuospatial ability, and the tasks were to mentally assemble fragmented geometric shapes to see which complete figure they would form. *Technical Comprehension* was used to test technical/mechanical ability, and the tasks were to solve problems related to basic physics and mechanics (e.g., gears, pulleys, and leverage) through illustrations.

## Mortality ascertainment

All-cause and cause-specific mortality was ascertained through linkage with the Swedish Cause of Death Register which provides complete populations coverage. Underlying causes of death were harmonized across ICD-7 to ICD-10. Dates of death were used to construct time-to-event outcomes. Deaths were grouped into mutually exclusive categories: cardiovascular, cancer, respiratory, external, and other. External causes were further subdivided into accidental, suicide, homicide, and undetermined intent based on standard ICD intent groupings. Details and full operational definitions are provided in the Supplementary Information.

## Statistical analysis

All analyses were conducted using Cox proportional hazards regression with attained age as the underlying time scale, specifying start and stop ages to account for delayed entry. Baseline hazards were stratified by birth cohort (1948 and 1953) to allow for cohort-specific age distributions and follow-up periods. The primary predictors were childhood general cognitive ability at age 13 years (child g) and adolescent cognitive change, operationalized as late-adolescent general cognitive ability residualized with respect to childhood cognitive level (army g residual). All cognitive predictors and parental education were standardized (per SD) within cohort, with the exception that residual g scores were not re-standardized (see above). Proportional hazards assumptions were evaluated using Schoenfeld residuals.

### Measurement-error calibrated sensitivity analysis

Because cognitive scores are measured with error, the residualized adolescent cognitive score may partly reflect stable cognitive trait variance not captured at age 13 rather than genuine adolescent change. We therefore conducted a secondary measurement-error calibrated analysis. We used a two-occasion classical measurement model in which latent cognitive ability at ages 13 and 18 years is decomposed into childhood level and true rank-order change. The model was anchored using the observed age-13 to age-18 cognitive correlation in the present data (r ≈ 0.77) and external meta-analytic estimates of reliability-corrected latent stability of general cognitive ability over a comparable interval (ρ= 0.891, 95% CI: 0.86-0.92) (33). Under the main anchor, latent stability was set to 0.89, yielding an implied reliability of approximately 0.85 and a latent-change SD of approximately 0.51 SD units. We then applied multivariate regression calibration to the two Cox coefficients for childhood cognitive level and adolescent residualized change. These calibrated estimates are reported as sensitivity analyses because they depend on measurement-model assumptions and external stability anchors.

Primary models jointly included childhood cognitive level and adolescent cognitive change to assess their independent associations with all-cause and cause-specific mortality. To evaluate robustness to early-life socioeconomic background, models were additionally adjusted for parental education. Hazard ratios (HRs) and 95% confidence intervals (CIs) are reported.

Cause-specific mortality analyses were conducted using the same modeling framework, with deaths grouped into mutually exclusive categories and external causes further subdivided. To complement analyses of individual causes, we examined a broader non-external (“intrinsic”) mortality category comprising cancer, cardiovascular, respiratory, and remaining non-external causes. Detailed operational definitions and supplementary analyses, including alternative operationalizations of cognitive change and full age-specific results, are provided in SI.

As an exploratory analysis motivated by the observed cause-specific patterns, we conducted a formal heterogeneity analysis contrasting cancer and accidental mortality. Using a stacked-data Cox approach, we tested whether associations of childhood cognitive level and adolescent cognitive change differed across these mortality categories by including interaction terms between cognitive predictors and cause group. This analysis was intended to evaluate whether descriptive differences observed in the primary cause-specific analyses were accompanied by evidence of coefficient heterogeneity.

## Data Availability

Individual-level data from the Evaluation Through Follow-up (UGU) study and linked Swedish national health registers are not publicly available because they are subject to Swedish and EU data protection legislation and ethical approvals that restrict data sharing. Researchers may apply for access to the UGU data through the University of Gothenburg and, where applicable, to linked Swedish national register data through the relevant Swedish authorities, subject to ethical approval and data protection agreements. Information about the application process for UGU data is available at: https://www.gu.se/en/evaluation-through-follow-up/disclosure-of-ugu-data.

## Acknowledgements

We thank all participants in the UGU 1948 and 1953 cohorts, Lucas Fischer Madsen for database management, Dan-Olof Rooth for generously helping with the army education codes, Anders M. Fjell for commenting on the manuscript, and Torstein Velsand for inspiring the analytic framework. Kristine B. Walhovd, affiliated with the University of Gothenburg, Sweden, and University of Oslo, Norway, conducted and is responsible for the data analysis. Kristine B. Walhovd had full access to all the data in the study and takes responsibility for the integrity of the data and the accuracy of the data analysis. Walhovd used ChatGPTversion 5.2 (OpenAI, San Francisco, CA, USA) and Claude Opus 4.6 (Anthropic, San Fransisco, CA, USA) to help shorten the abstract and significance statement and part of the introduction, and to help debug part of the code written by Walhovd. Walhovd takes responsibility for the integrity of the content thus revised by this tool and revised all contents herself. Additionally, A. Segerberg also manually inspected the code. All authors took part in concept and design and undertook critical revision of the manuscript for important intellectual content.

