## Supplementary Information for "Change for life? Adolescent cognitive development predicts mortality risk independent of childhood ability"

for

Walhovd et al.

**Content:**

**Supplementary results**	p 3

Figure S1 Age distribution of deaths by cause in the analytic sample	p 3

Analyses using childhood and adolescent cognitive ability	p 4

Analyses using delta cognitive change scores (Δg)	p 4

Decomposition and measurement-error calibration, residualized measure	p 5

Figure S2. Reliability-bounded interpretation, residualized change	p 7

Figure S3. Observed-score and measurement-error calibrated associations. p 9

Exploratory heterogeneity analyses: cancer versus accidental mortality	p 10

Analyses considering possible loss-to-follow up by migration	p 10

**Table S1. Correlates of adolescent cognitive change	p 11**

Table S2A. Cox model of all-cause mortality, 1948 cohort	p 13

Table S2B. Cox model of all-cause mortality,1948 cohort, adjusted	p 13

Table S3A. Cox model of accidental mortality, 1948 cohort	p 14

Table S3B. Cox model of accidental mortality,1948 cohort, adjusted	p 14

Table S4A. Cox model of all-cause mortality, 1953 cohort	p 15

Table S4B. Cox model of all-cause mortality,1953 cohort, adjusted	p 15

Table S5A. Cox model of accidental mortality, 1953 cohort	p 16

Table S5B. Cox model of accidental mortality,1953 cohort, adjusted	p 16

**Supplementary methods**	p 17

Figure S4. Sample selection flowchart for the UGU 1948 and 1953 cohorts. p 17

Sample selectivity analyses	p 18

Socioeconomic and psychosocial measures	p 20

Classification of cause-specific mortality	p 22

**References**	p 26

**Supplementary Results**

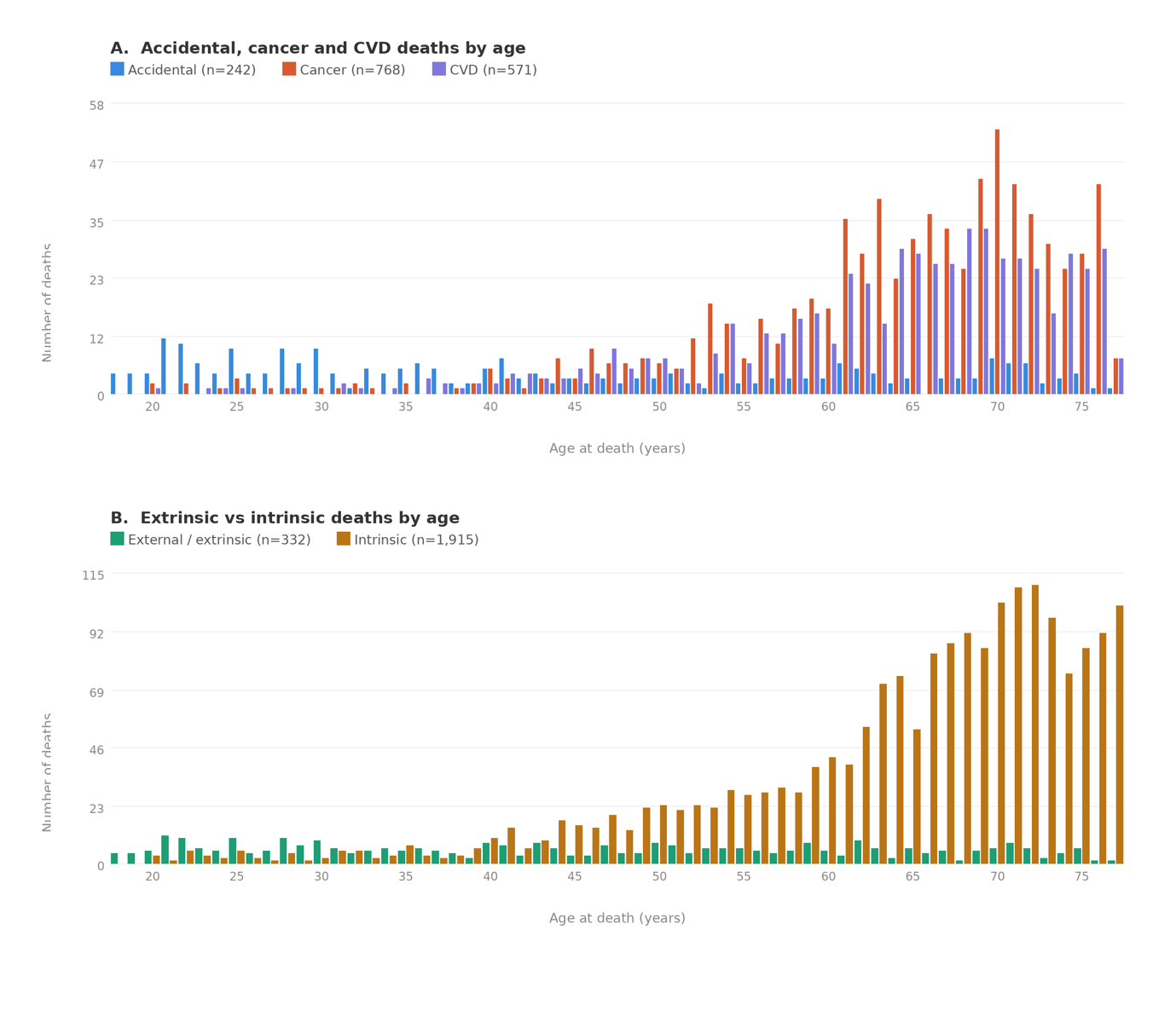
***Figure S1. Age distribution of deaths by cause in the analytic sample.***

*Panel A shows the number of deaths per year of age in the analytic sample (n = 9412) for accidental (n = 242), cancer (n = 768), and cardiovascular (n = 571) causes. Panel B shows deaths by year of age for extrinsic (external causes, n = 332) and intrinsic causes (cancer, cardiovascular, respiratory, and other, n = 1,915). Extrinsic deaths are proportionally more frequent in early adulthood, while intrinsic deaths increase steeply from midlife onward, with the crossover occurring around age 50.*

Proportional hazards assumptions for the primary all-cause mortality model including childhood cognitive level, adolescent cognitive change, and parental education were evaluated using Schoenfeld residuals. No evidence of substantial violation was observed for individual predictors (childhood cognitive level: p = .130; adolescent cognitive change: p = .070; parental education: p = .745) or globally (p = .097).

***Analyses using childhood and adolescent cognitive ability***

When childhood and adolescent cognitive ability were included jointly, both remained independently associated with mortality risk, although effect sizes were attenuated. A one–standard deviation higher childhood cognitive ability was associated with an approximately 8% lower mortality risk (HR = 0.92; 95% CI, 0.87–0.98), while the same for adolescent cognitive ability was associated with a 17% lower mortality risk (HR = 0.83; 95% CI, 0.78–0.88), indicating that both cognitive performance in childhood and late adolescence provides predictive information beyond either measure alone. Adjustment for parental education modestly attenuated effect estimates but did not alter the pattern of results. Both childhood cognitive ability (HR = 0.93; 95% CI, 0.87–0.99) and adolescent cognitive ability (HR = 0.84; 95% CI, 0.79–0.89) remained independently associated with mortality risk, while higher parental education was itself associated with lower mortality (HR per SD = 0.95; 95% CI, 0.90–1.00, p = .0356).

***Analyses using delta cognitive change scores (Δg)***

To assess whether results depended on the operationalization of cognitive change across adolescence, we conducted supplementary analyses using a simple difference score (Δg; late-adolescent minus childhood general cognitive ability) in place of the residualized adolescent cognitive change measure used in the main analyses. When childhood cognitive level is included in the model, Δg and residualized change are identified from the same variation in adolescent cognition independent of childhood level, and therefore yield equivalent estimates for the developmental component in linear models. However, Δg retains variance shared with childhood cognitive level and thus reflects a mixture of developmental change and stable individual differences already present in childhood. In models including childhood cognitive level and Δg, both predictors were independently associated: A one–standard deviation increase in childhood cognitive level was associated with a lower mortality risk (HR = 0.77; 95% CI, 0.74–0.80; p < 2×10⁻¹⁶), and higher Δg was similarly associated with reduced mortality risk (HR = 0.88; 95% CI, 0.84–0.92; p = 5.5×10⁻⁹). Effect estimates for childhood cognitive level were stronger than in corresponding models using residualized adolescent cognitive change, consistent with Δg retaining variance shared with childhood cognitive ability. For this reason, residualized adolescent cognitive change was retained as the primary operationalization of cognitive development across adolescence in the main analyses, as it more cleanly separates baseline cognitive level from developmental divergence.

***Decomposition and measurement-error calibration of the residualized adolescent cognitive measure***

A potential concern with interpreting the residualized adolescent cognitive score as developmental change is that, under classical measurement error, the residual may partly reflect stable cognitive ability not captured by the childhood measure, rather than genuine adolescent cognitive change. To evaluate this possibility, we applied a two-occasion classical measurement model decomposing the residualized age-18 cognitive score into genuine rank-order change, stable-trait recovery from imperfect childhood measurement, and measurement noise.

Let $g_{1}$ denote latent cognitive ability at age 13 and $g_{2}$ latent cognitive ability at age 18, with

$$g_{2}=g_{1}+\delta,$$

where $\delta$ represents true rank-order change across adolescence. Observed cognitive scores were modeled as noisy indicators of latent ability,

$$T_{i}=\lambda g_{i}+\epsilon_{i},$$

with $\lambda^{2}=\rho^{2}$ representing effective reliability and $\epsilon_{i}$ classical measurement error. Scores were standardized within wave, matching the procedure used in the main analyses. Under this model, the observed age-13 to age-18 correlation reflects both latent stability and measurement error.

We anchored the model using the observed correlation between childhood and late-adolescent cognitive ability in the present sample, $r\approx.77$, together with external meta-analytic estimates of reliability-corrected latent stability of general cognitive ability across a comparable interval. Breit et al. (1) reported a reliability-adjusted latent stability for general cognitive ability of approximately $c=.89$. Combining this anchor with the observed correlation yielded an implied effective reliability of approximately $\rho^{2}=.85$ and a latent-change SD of approximately 0.51 latent-g units, corresponding to about 8 rank-order IQ points (Figure S2).

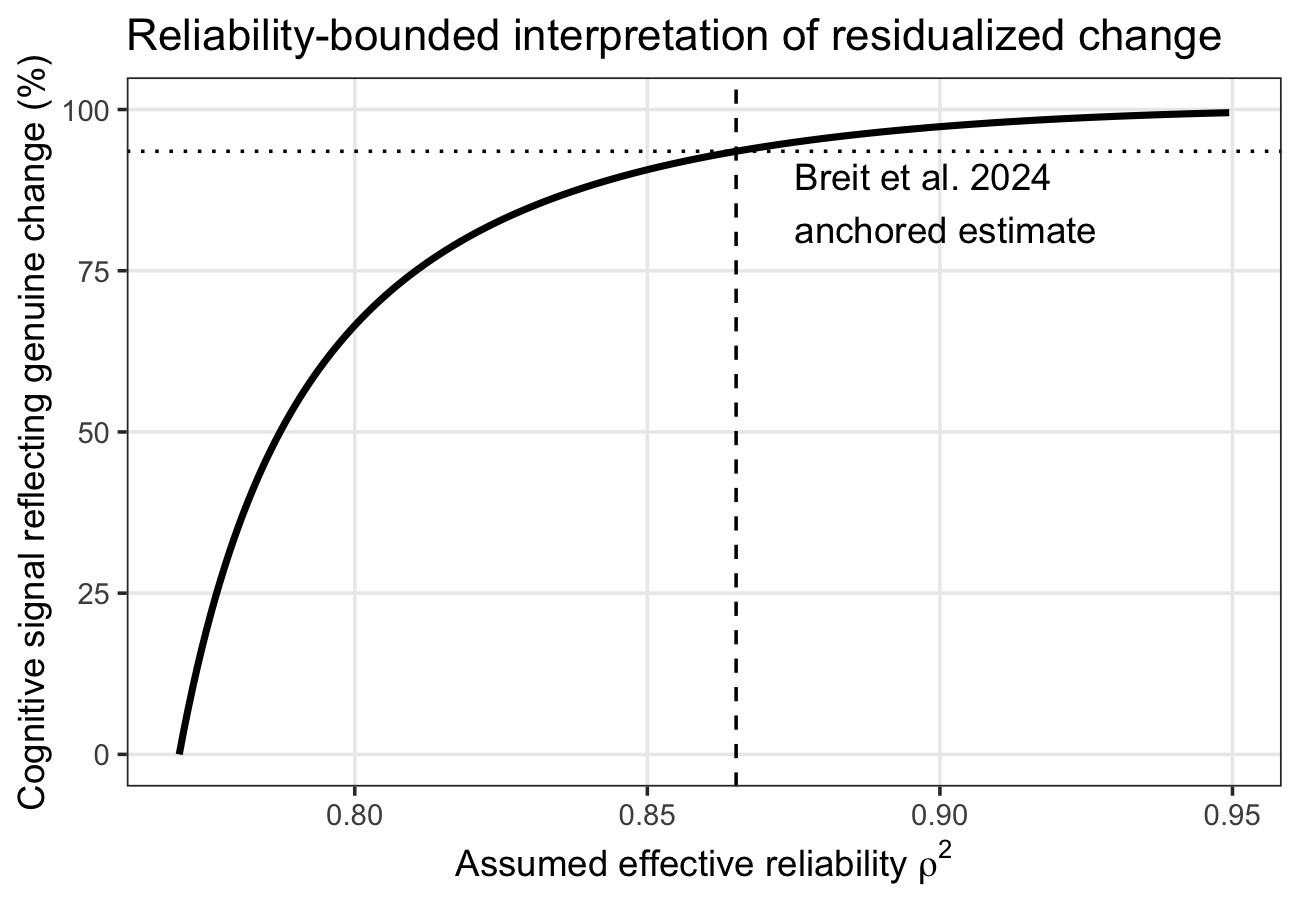

***Figure S2. Reliability-bounded interpretation of residualized adolescent cognitive change***

*Estimated proportion of the cognitive signal in the residualized adolescent cognitive score attributable to genuine adolescent cognitive change under varying assumptions about test reliability. The dashed vertical line indicates the reliability estimate implied by combining the observed age-13 to age-18 cognitive correlation in the present data (*$r\approx.77$*) with an external latent-stability anchor for general cognitive ability (*$c\approx.89$*). At this anchor, approximately 92% of the cognitive signal reflects genuine adolescent change rather than stable-trait recovery from imperfect childhood measurement.*

Under these assumptions, approximately 92% of the cognitive signal in the residualized adolescent score reflected genuine adolescent change, whereas approximately 8% reflected stable-trait variance not captured by the childhood measure. Of the total residualized-score variance, approximately 45% reflected genuine change, 4% stable-trait recovery, and 51% classical measurement noise. Thus, the residualized adolescent score is expected to be attenuated by measurement noise, but its cognitive signal is dominated by genuine developmental divergence rather than recovery of stable trait variance. Across plausible latent-stability values spanning the confidence interval of the external anchor, conclusions remained essentially unchanged: the estimated genuine-change share remained high and the interpretation of the residualized adolescent measure as primarily reflecting developmental divergence was preserved.

Because the mortality models include both childhood cognitive level and residualized adolescent change, scalar reliability correction is not appropriate. Measurement error in one covariate can bias coefficients for both covariates, requiring a multivariate correction. We therefore applied regression calibration using the matrix correction implied by the same measurement model. For the un-restandardized residual used in the main analyses, evaluated at the externally anchored point, the correction matrix was

$$K=\left( \begin{matrix} 1.085 & 0 \\ -0.626 & 1.882 \end{matrix} \right).$$

This matrix maps the observed Cox coefficient vector for childhood cognitive level and adolescent residualized change to latent-scale coefficients for childhood cognitive level and true adolescent change. Corrected estimates are reported as sensitivity analyses, not replacements for the primary observed-score Cox estimates, because they depend on assumptions about latent stability, measurement structure, and classical measurement error (Figure S3).

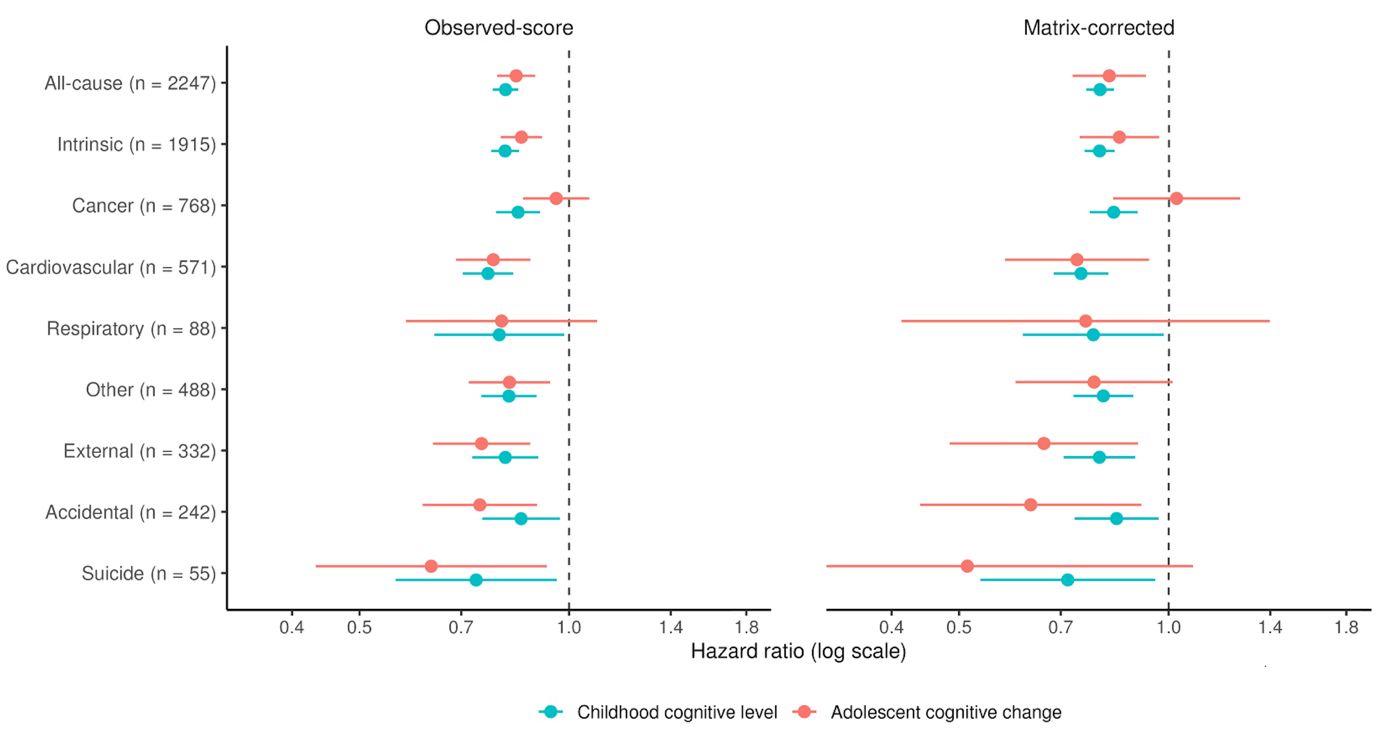

***Figure S3. Observed-score and measurement-error calibrated mortality associations***

*Raw/ Observed-score Cox estimates are the primary estimates as reported in the manuscript. Matrix-corrected estimates are secondary regression-calibrated estimates under a two-occasion classical measurement-error model anchored to external estimates of latent cognitive stability. Corrected estimates are expressed per latent-g SD and are intended as sensitivity analyses, not replacements for the primary observed-score estimates.*

Simulation checks under the same measurement model indicated that the matrix correction recovered latent parameters more accurately than either uncorrected Cox estimates or scalar reliability corrections. The measurement-error calibrated estimates sharpened the main cause-specific pattern: associations for external and accidental mortality strengthened, whereas the already weak adolescent-change association for cancer mortality attenuated further.

**Exploratory heterogeneity analyses: cancer versus accidental mortality**

To formally evaluate whether the descriptive cause-specific patterns reflected differences in coefficient magnitude, we conducted exploratory stacked-data Cox analyses comparing cancer and accidental mortality. Cancer and accidental deaths were selected because they represented the clearest contrast in the primary analyses, with childhood cognitive level showing stronger associations with cancer mortality and adolescent cognitive change showing stronger associations with accidental mortality, and this pattern was also further emphasized in measurement error calibrated mortality associations. Formal heterogeneity analyses indicated that associations of adolescent cognitive change differed across mortality categories (χ² = 5.42, df = 1, p = .020), whereas childhood cognitive level did not (χ² = 0.08, df = 1, p = .78). The joint test across both cognitive predictors approached significance (χ² = 5.55, df = 2, p = .062). These findings paralleled the descriptive patterns observed in the primary analyses and suggest that developmental divergence across adolescence may be relatively more strongly linked to accidental than cancer mortality.

***Sensitivity analyses considering possible loss-to-follow up by migration***

Sensitivity analyses censoring follow-up at last recorded emigration among individuals with emigration without return (n = 164) yielded materially unchanged results compared to the primary analyses on all-cause mortality (childhood cognitive ability: HR, 0.81; 95% CI, 0.77–0.84; adolescent cognitive change: HR, 0.83; 95% CI, 0.78–0.89), indicating that selective emigration did not explain the observed associations.

| **A. 1948 cohort variables** | **Estimate (SE)** | **p** |
| --- | --- | --- |
| Intercept | 0.159 (0.065) | 0.014 |
| Childhood cognitive level (z) | -0.038 (0.009) | <0.001 |
| Parental education (z) | 0.071 (0.018) | <0.001 |
| Paternal occupation: higher officials/business | 0.032 (0.054) | 0.547 |
| Paternal occupation: other officials/business | 0.033 (0.070) | 0.639 |
| Paternal occupation: farmers | -0.196 (0.072) | 0.006 |
| Paternal occupation: workers | -0.091 (0.070) | 0.190 |
| Distance to secondary school | -0.050 (0.010) | <0.001 |
| **B. 1953 cohort variables** |  |  |
| Intercept | -0.073 (0.064) | 0.258 |
| Childhood cognitive level (z) | -0.084 (0.010) | <0.001 |
| Parental education (z) | 0.079 (0.010) | <0.001 |
| Education at conscription | 0.132 (0.011) | <0.001 |
| Non-psychiatric diagnosis at conscription | -0.040 (0.021) | 0.054 |
| Psychiatric non-substance diagnosis | -0.270 (0.028) | <0.001 |
| Substance abuse diagnosis at conscription | -0.413 (0.076) | <0.001 |
| Vision | -0.018 (0.007) | 0.006 |

**Table S1. Correlates of adolescent cognitive change in the 1948 and 1953 cohorts.**
Linear regression models predicting adolescent cognitive change from childhood cognitive level and cohort-specific contextual, educational, and health-related factors collectively. Outcome was adolescent cognitive change, operationalized as late-adolescent general cognitive ability residualized with respect to childhood cognitive level. Paternal occupation was modeled categorically, with the highest occupational group (acedemics/professionals) as reference. Because adolescent cognitive change was operationalized as late-adolescent cognitive ability residualized with respect to childhood cognitive level, childhood cognitive level was included as a covariate in models for consistency across the analytic subsamples used in each cohort-specific analysis. Small remaining associations reflect sample restriction and covariate missingness rather than substantive dependence of the residualized outcome on childhood level. Cohort-specific predictors reflected variables available in each cohort (df= 4771 in 1948, 4345 in 1953), see SI methods.

| **Variable** | **HR (95% CI)** | **p** |
| --- | --- | --- |
| Childhood cognitive level (z) | 0.81 (0.77–0.85) | >0.001 |
| Adolescent cognitive change (residual g) | 0.86 (0.80–0.93) | >0.001 |

**Table S2A. Cox proportional hazards model of all-cause mortality in the 1948 cohort, unadjusted** (n = 5,001; deaths: 1416)**.**

| **Variable** | **HR (95% CI)** | **p** |
| --- | --- | --- |
| Childhood cognitive level (z) | 0.82 (0.77–0.86) | >0.001 |
| Adolescent cognitive change | 0.88 (0.81–0.95) | 0.002 |
| Parental education (z) | 0.93 (0.82–1.05) | 0.225 |
| Distance to secondary school | 0.99 (0.94–1.05) | 0.772 |
| Paternal occupation: higher officials/business | 1.03 (0.71–1.49) | 0.883 |
| Paternal occupation: other officials/business | 0.99 (0.63–1.57) | 0.974 |
| Paternal occupation: farmers | 0.95 (0.59–1.53) | 0.834 |
| Paternal occupation: workers | 1.18 (0.75–1.87) | 0.474 |

**Table S2B. Cox proportional hazards model of all-cause mortality in the 1948 cohort adjusted for parental education, distance to school, and paternal occupational class**. n = 4,779; deaths = 1340.

| **Variable** | **HR (95% CI)** | **p** |
| --- | --- | --- |
| Childhood cognitive level (z) | 0.83 (0.70–0.99) | 0.034 |
| Adolescent cognitive change (residual g) | 0.74 (0.58–0.96) | 0.022 |

**Table S3A. Cox proportional hazards model of accidental mortality in the 1948 cohort, unadjusted** (n = 5,001; accidental deaths: 129)**.**

| **Variable** | **HR (95% CI)** | **p** |
| --- | --- | --- |
| Childhood cognitive level (z) | 0.85 (0.71–1.02) | 0.080 |
| Adolescent cognitive change | 0.74 (0.57–0.98) | 0.033 |
| Parental education (z) | 1.26 (0.86–1.86) | 0.233 |
| Distance to secondary school | 1.00 (0.83–1.21) | 0.977 |
| Paternal occupation: higher officials/business | 0.99 (0.29–3.33) | 0.987 |
| Paternal occupation: other officials/business | 1.54 (0.31–7.61) | 0.597 |
| Paternal occupation: farmers | 1.59 (0.31–8.16) | 0.581 |
| Paternal occupation: workers | 2.37 (0.48–11.64) | 0.288 |

**Table S3B. Cox proportional hazards model of accidental mortality in the 1948 cohort adjusted for parental education, distance to school, and paternal occupational class**. n = 4,779; accidental deaths = 120.

| **Variable** | **HR (95% CI)** | **p** |
| --- | --- | --- |
| **Childhood cognitive ability (z)** | 0.79 (0.74–0.85) | >0.001 |
| **Adolescent cognitive change (residual g)** | 0.78 (0.71–0.87) | >0.001 |

**Table S4A. Cox proportional hazards model of all-cause mortality in the 1953 cohort, unadjusted** (n = 4,411; deaths: 831).

| **Variable** | **HR (95% CI)** | **p** |
| --- | --- | --- |
| Childhood cognitive level (z) | 0.82 (0.76–0.88) | >0.001 |
| Adolescent cognitive change | 0.82 (0.74–0.91) | >.001 |
| Parental education (z) | 1.04 (0.96–1.12) | 0.315 |
| Education at conscription | 0.93 (0.86–1.01) | 0.086 |
| Non-psychiatric diagnosis at conscription | 1.02 (0.87–1.19) | 0.814 |
| Psychiatric non-substance diagnosis | 1.28 (1.05–1.55) | 0.015 |
| Substance abuse diagnosis at conscription | 1.86 (1.22–1.84) | 0.004 |
| Vision | 0.97 (0.93–1.02) | 0.282 |

**Table S4B. 1953 cohort: all-cause mortality adjusted for parental education, education at conscription, presence of any diagnosis and psychiatric diagnosis at conscriptiion** (n = 4,353; deaths = 818).

| **Variable** | **HR (95% CI)** | **p** |
| --- | --- | --- |
| **Childhood cognitive ability (z)** | 0.86 (0.72–1.04) | 0.127 |
| **Adolescent cognitive change (residual g)** | 0.74 (0.56–0.97) | 0.032 |

**Table S5A. Cox proportional hazards model of accidental mortality in the 1953 cohort, unadjusted** (n = 4,411; accidental deaths: 113).

| **Variable** | **HR (95% CI)** | **p** |
| --- | --- | --- |
| Childhood cognitive level (z) | 0.86 (0.71–1.06) | 0.152 |
| Adolescent cognitive change | 0.77 (0.58–1.03) | 0.079 |
| Parental education (z) | 0.92 (0.73–1.15) | 0.466 |
| Education at conscription | 1.18 (0.95–1.47) | 0.145 |
| Non-psychiatric diagnosis at conscription | 1.12 (0.73–1.71) | 0.597 |
| Psychiatric non-substance diagnosis | 1.28 (0.73–2.22) | 0.387 |
| Substance abuse diagnosis at conscription | 2.54 (0.89–7.26) | 0.082 |
| Vision | 0.95 (0.86–1.09) | 0.516 |

**Table S5B. 1953 cohort: accidental mortality adjusted for parental education, education at conscription, presence of any diagnosis and psychiatric diagnosis at conscriptiion** (n = 4,353; accidental deaths = 111).

**Supplementary methods**

**Sampling frame**

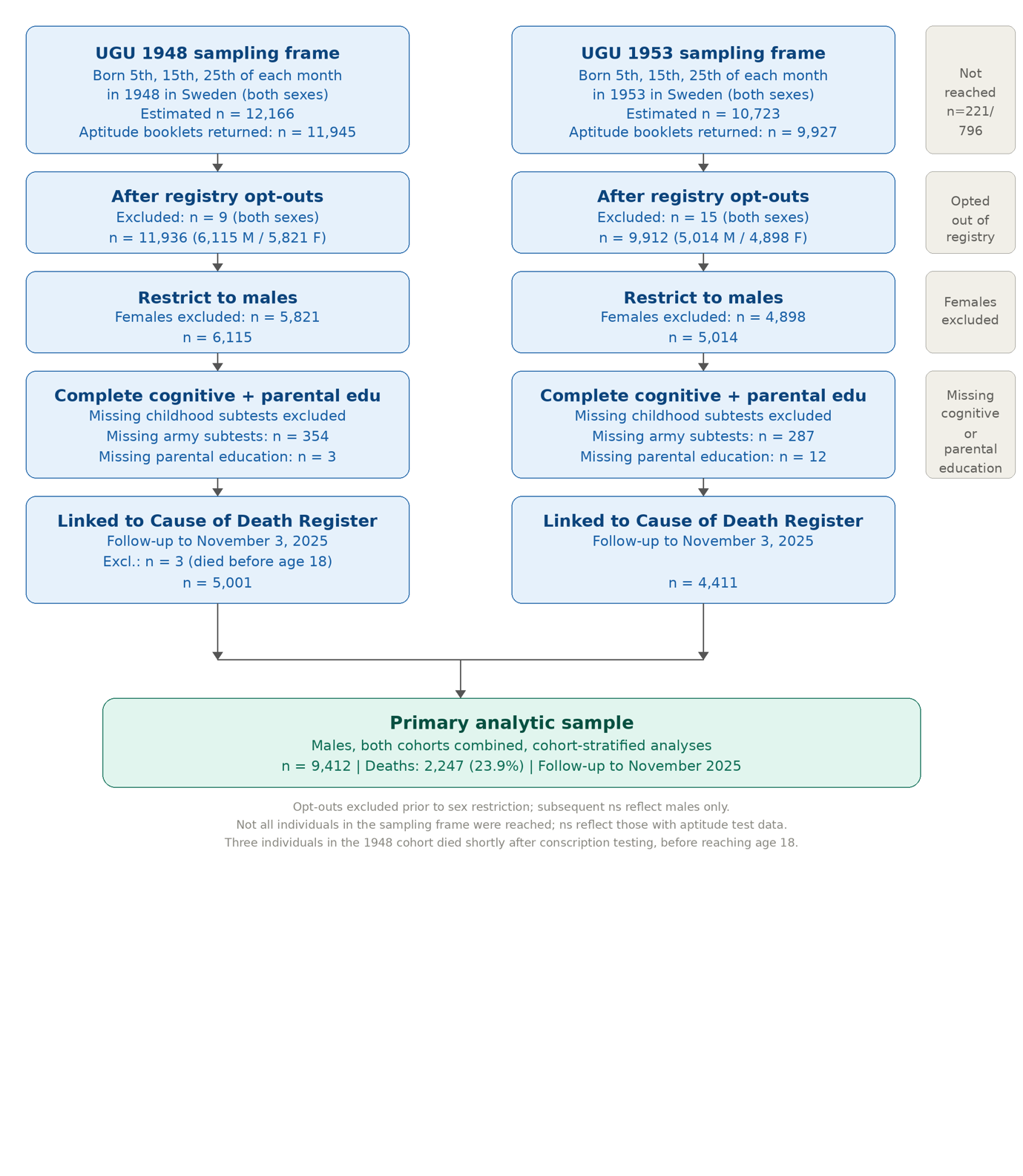

**Figure S4. Sample selection flowchart for the UGU 1948 and 1953 cohorts.**

The UGU cohorts comprise nationally representative samples of individuals born on the 5th, 15th, and 25th of each month in 1948 and 1953 in Sweden. Not all individuals in the sampling frame were reached at the time of data collection; ns in the top boxes reflect those from whom aptitude test booklets were returned. Individuals who opted out of registry data extraction were excluded prior to sex restriction. Subsequent sample sizes reflect males only, as the present study is restricted to males due to the requirement of military conscription data. Males with missing data on any army cognitive subtest, any childhood cognitive subtest, or parental education, or not alive at age 18, were excluded. The primary analytic sample was linked to the Swedish Cause of Death Register with follow-up through November 3, 2025. M = male; F = female.

***Sample selectivity analyses***

Twenty-four persons, of whom ten who otherwise would have been eligible for inclusion in the analytic sample since they had data on all subtests at age 13 and 18 and parental education, noted that they did not want to have further registry data extracted in connection with the study, and they were not included in analyses. This sample was considered too small for selectivity analyses based on childhood data.

While the army conscription testing was mandatory for all Swedish males of the relevant birth cohorts, conscription data do still not cover the full population, and a minority with certain health conditions of relevance to later mortality were exempt (2). Complete army cognitive data were not available for 354 children in the 1948 cohort, and 287 children in the 1953 cohort who otherwise had measures of all childhood cognitive tests and parental education. Three cases in the 1948 and 12 in the 1953 with complete cognitive data across timepoinst lacked a measure of parental education.

To assess potential selection into the subsample with conscription-based cognitive data, we examined whether childhood cognitive ability sum score (TS6STP) predicted availability of complete army cognitive test scores. In the 1948 cohort, childhood cognitive ability was not significantly associated with having complete army data, but a negative trend was observed, meaning higher ability children tended to less likely have army cognitive tests (β = −0.006, SE = 0.003, p = 0.064). In the 1953 cohort, higher childhood cognitive ability was associated with an increased likelihood of having complete army data (β = 0.019, SE = 0.003, p < 0.001). In pooled analyses across cohorts, higher childhood cognitive ability (sum score) was significantly associated with a increased likelihood of having complete army data, though the effect was not large (β = 0.006, SE = 0.002, p = 0.009). Thus, evidence for selective inclusion based on childhood cognitive ability was modest and not consistent across cohorts. Such selectivity would most likely bias estimates toward the null by slightly restricting the range of cognitive ability in the analytic sample. Thus, the observed associations between cognitive ability and mortality are unlikely to be explained by selective inclusion and may, if anything, be conservative. However, this should be noted as a limitation.

We did not exclude persons based on possible emigration, since the Swedish CDR aims to include, and does include, also information on deceased persons not registered in Sweden (3). Still, to evaluate potential bias from possible incomplete follow-up due to emigration, we conducted selectivity and sensitivity analyses using migration register data. Individuals could have multiple migration records, including emigration and later immigration. We therefore classified migration status based on the last recorded migration event. In three cases, migration-based censoring dates preceded the analytic start of follow-up despite observed participation in the conscription-based data. These cases were considered likely to reflect anomalous migration records (4) and were excluded from the migration sensitivity analysis. In sensitivity analyses, participants whose last recorded migration was emigration and who had no recorded death were censored at their last emigration date rather than followed until the end of register follow-up. This affected 164 individuals in the analytic sample, leaving 9409 individuals and 2247 deaths in main migration sensitivity analyses.

We examined whether childhood cognitive ability and adolescent cognitive change predicted emigration without recorded return, to assess the potential for selective loss to follow-up. We fitted a logistic regression model predicting emigration without return. Both higher childhood cognitive ability (OR, 1.91; 95% CI, 1.58–2.30) and greater adolescent cognitive change (OR, 1.75; 95% CI, 1.36–2.25) were associated with increased likelihood of emigration without return, indicating selective migration by cognitive ability.

We thus re-estimated the primary Cox proportional hazards models using attained age as the time scale, replacing the original follow-up time with migration-adjusted follow-up (stop_age_sens), defined as the minimum of the original end of follow-up and age at last recorded emigration among individuals with emigration without return and no recorded death. Models included childhood cognitive ability and adolescent cognitive change simultaneously and were stratified by cohort (see Supplementary results).

***Socioeconomic and psychosocial measures***

Parental education was recorded at the time of testing in 6th grade of primary school as a school administrative variable for the 1948 cohort, and by questionaires to students in the 1953 cohort, and coded on an ascending scale from 1 to 4. The mean of paternal and maternal education, or either parent´s education if one was missing, was used in analyses (M = 1.259, SD = 0.574). Of note, for 77%, this value was 1, indicating only elementary parental education.

For the 1948 cohort, school adminstrative information was also available for: Father´s occupation grouped as a categorical variable (n = 4784): 5.6% A: akademics, officers, teachers, business people in leading positions, 9.7% B: officials, businessmen, with lower school certificate or higher theoretical educaction; 21.0% C: other officials and businessmen; 14.4% D: Farmers, and 49.3% E: Worker. Length of route between home at different types of schools namely compulsory, junior secondary (n = 4996), and upper secondary school (n = 4992), coded as a numeric variable from 1 (less than 20 minutes), 2 (20-40 minutes), 3 (40-60 minutes), 4 (60-80 minutes), to 5 (more than 80 minutes). 46.0 % lived within 20 minutes from junior secondary school, 34.6% within 40 minutes, 11.5% within 60 minutes, 4.7% within 80 minutes, and 3.1% more than 80 minutes from junior secondary school. The correspoding numbers for upper secondary school were 36.6, 31,4%, 13.7%, 7,7%, and 10.5%. We used distance from junior secondary school in analyses of correlates of cognitive change.

For the 1953 cohort, detailed education and health data were available from the digital conscription archive, including conscription files from 1969 onwards (2). For the present analyses, data were extracted for education and presence of medical diagnosis. Education level was classified on a 7-level scale reflecting the highest completed level of formal education (n = 4355), 34.9% had pre-upper secondary education of ≤9 years, and 14.6% had exactly 9 years of pre-upper secondary education. Upper secondary education of up to 2 years was completed by 49,5%, whereas 0.8% had completed longer upper secondary education (>2 and ≤3 years). 4 persons were listed as having completed short post-secondary education (<3 years, including 4-year upper secondary programs), and 3 with long post-secondary education (≥3 years). Doctoral education was not present in the sample. Presence of any medical diagnosis was coded as 0 and 1, with 2459 instances of at least one medical diagnosis. Whether a main diagnosis was a psychiatric disorder, defined as ICD 8 codes 290-315 (5), was coded as 1 versus 0 if other or no diagnosis, with 358 instances of a psychiatric diagnosis, and across main and side diagnoses (a maximum of 6, including main), 734 had a psychiatric diagnosis, of which only 4 involved psychotic diseases (ICD 295-298). None had hyperkinetic disorder (ADHD). A diagnosis involving substance abuse (i.e. ICD 8 codes 291, 303,304) was given in 70 cases. For the analyses involving diagnostic variables at conscription, we coded the diagnostic classes as mutually exclusive, to enter them simultanously in analyses. Vision, or eyesight, as graded at conscription on a variable from 1-9 (n= 4406, mean: 8.14, SD = 1.42) was also included in analysis.

***Classification of cause-specific mortality***

Underlying cause of death was obtained from the Swedish Cause of Death Register (CDR) using the variable ULORSAK, which records the underlying cause according to the ICD revision in use at the time of death (ICD-7, ICD-8, ICD-9, or ICD-10). In a minority of cases, exact date is missing from CDR (3). If exact day of death was missing this was set to the first of the month (n = 142), and if month of death was missing (n = 22), this was set to the first month of the year of death. Because ICD code formats and cause definitions differ across revisions, we implemented a harmonized classification procedure to assign each death to one mutually exclusive and interpretable cause-of-death category. Cardiovascular deaths included diseases of the circulatory system (ICD-10 I codes and corresponding ICD-7/8/9 ranges), cancer deaths included malignant neoplasms (ICD-10 C codes; ICD-7/8/9 140–299), and respiratory deaths included diseases of the respiratory system (ICD-10 J codes and corresponding ICD-7/8/9 ranges). External causes comprised injuries and violence (ICD-10 V–Y; ICD-7/8/9 800–999) and were further subdivided into accidental, suicide, homicide, and undetermined intent based on standard ICD intent groupings (e.g., ICD-10 X60–X84 for suicide, X85–X99 for homicide, and Y10–Y34 for undetermined intent).

All cause-of-death codes were converted to upper case and trimmed of whitespace. For ICD-10, chapter letters were identified from the first character of the code. For ICD-7/8/9, numeric codes were identified by extracting the first three digits. For external causes recorded using E-codes or extended numeric formats, numeric components were extracted as needed to ensure consistent classification across ICD revisions. Deaths were first classified into broad cause-of-death groups (cardiovascular, cancer, respiratory, external, other). Deaths classified as external causes were subsequently subdivided by intent into accidental, suicide, homicide, or undetermined intent, as described below.

Deaths were classified as *cardiovascular* if the underlying cause corresponded to diseases of the circulatory system, including cerebrovascular disease, defined as: ICD-10 codes beginning with I; ICD-8/9 three-digit codes 390–459; and ICD-7 circulatory diseases (400–468) and cerebrovascular diseases (330–334). This category captures major fatal cardiovascular and cerebrovascular conditions across ICD revisions. Deaths were classified as *cancer* if the underlying cause corresponded to malignant neoplasms, defined as: ICD-10 codes beginning with C; ICD-7/8/9 three-digit codes 140–209. Cancer deaths were grouped irrespective of tumor site or metastatic status. Deaths were classified as *respiratory* if the underlying cause corresponded to diseases of the respiratory system, defined as: ICD-10 codes beginning with J; ICD-8/9 three-digit codes 460–519; and ICD-7 three-digit codes 470–527.

Deaths were classified as *external causes* if the underlying cause corresponded to injury, poisoning, or violence, defined as: ICD-10 external-cause chapters V–Y; ICD-7/8/9 three-digit codes 800–999, including E-codes with numeric ranges 800–999 where applicable. External causes were subsequently subdivided by intent as described below.

All remaining deaths not meeting criteria for the above categories were classified as *other* causes. To complement analyses of individual causes, we examined a broader non-external ("intrinsic") mortality category comprising cancer, cardiovascular, respiratory, and remaining non-external (“other”) causes.

*External causes by intent*

Deaths classified as external causes were further subdivided into mutually exclusive intent categories using ICD-revision-specific code ranges.

Deaths were classified as *suicide* if the underlying cause indicated intentional self-harm, defined as: ICD-10 codes X60–X84; ICD-8/9 E-codes E950–E959; and ICD-7 E-codes E970–E979. Numeric representations of these code ranges were used where applicable in early registry data. Deaths were classified as *homicide* if the underlying cause indicated assault by another person, defined as: ICD-10 codes X85–Y09; ICD-8/9 E-codes E960–E969; and ICD-7 E-codes E980–E985. Deaths were classified as *undetermined intent* if the underlying cause reflected uncertainty regarding intent, defined as: ICD-10 codes Y10–Y34 and ICD-8/9 E-codes E980–E989. No category for undetermined intent exists in ICD-7; such deaths were therefore not classified as undetermined for ICD-7. Deaths were classified as *accidental* if they met criteria for external causes but did not meet criteria for suicide, homicide, or undetermined intent. This category therefore includes unintentional injuries such as transport accidents, falls, poisonings, and other accidental events. External-cause subcategories were assigned hierarchically to ensure mutual exclusivity: suicide, homicide, and undetermined intent were identified first, and all remaining external deaths were classified as accidental.
